# Analysis of Fatality Impact and Seroprevalence Surveys in a Community Sustaining a SARS-CoV-2 Superspreading Event

**DOI:** 10.1101/2022.01.26.22269805

**Authors:** Enrico Richter, Dominik Liebl, Bianca Schulte, Nils Lehmann, Christine Fuhrmann, Karl-Heinz Jöckel, John P.A. Ioannidis, Hendrik Streeck

**Affiliations:** Institute of Virology, University Hospital, University of Bonn, Germany and German Center for Infection Research (DZIF), partner site Bonn-Cologne; Institute of Finance and Statistics and Hausdorff Center for Mathematics, University of Bonn, Germany; Institute of Medical Informatics, Biometry und Epidemiology (IMIBE), University Hospital Essen, Germany; Clinical Study Core Unit, Study Center Bonn (SZB), Institute of Clinical Chemistry and Clinical Pharmacology, University Hospital Bonn, Germany; Departments of Medicine, of Epidemiology and Population Health, of Biomedical Data Science, and of Statistics, Stanford University, Stanford, USA

**Author notes:** Corresponding author: Hendrik Streeck, Institute of Virology, University Hospital Bonn, Venusberg-Campus 1, 53127 Bonn, Germany.

## Abstract

There is ongoing debate on the COVID-19 infection fatality rate (IFR) and the impact of COVID-19 on overall population mortality. Here, we addressed these issues in a community in Germany with a major superspreader event analyzing deaths over time as well as auditing death certificates in the community.18 deaths that occurred within the first 6 months of the pandemic in the community had a positive test for SARS-CoV-2. Six out of 18 SARS-CoV-2+ deaths had non-COVID-19 related causes of death (COD). Individuals with confirmed infection and COVID-19 COD typically died of respiratory failure (75%) and tended to have fewer reported comorbidities (p=0.029). Duration between first confirmed infection and death was negatively associated to COVID-19 being COD (p=0.04). Repeated seroprevalence essays on an original sample of 587 individuals in three visits showed modest increases in seroprevalence over time, and substantial seroreversion (30% [27/90] (95% CI: [20.5%; 39.5%])). IFR estimates accordingly varied depending on COVID-19 death attribution and seroprevalence caveats. Careful ascertainment and audit of COVID-19 deaths are important in understanding the impact of the pandemic.

## Introduction

The COVID-19 pandemic has been associated with considerable mortality at a global level, but estimates about the total number of deaths have varied widely across populations, demographics and countries^1-5^. There is ongoing debate on the population specific COVID-19 fatality rates (IFRs) and on the impact of COVID-19 on overall population mortality in various settings^6^.

The IFR would be a robust marker for the severity of a virus-induced disease if the number of infected individuals (IFR denominator) as well as the number of deaths (IFR numerator) could be correctly measured. The number of infected individuals can be inferred by seroprevalence studies, and many such studies have been done^7,8^. However, biases often exist in such studies. Two main concerns are whether the surveyed sample is representative of the general population and whether people who get infected do not develop or lose their detectable antibodies over time. The validation of the number of COVID-19-related deaths has received less attention, but both over- and under-counting may occur^9^. COVID-19 deaths may have been underestimated in particular in some locations at the beginning of the pandemic when testing was limited, or because determining the cause of death (COD) in the elderly or those with multiple comorbidities was difficult. Conversely, COVID-19 deaths may have been over-attributed, e.g. if based on a positive SARS-CoV-2 test without a relevant clinical picture. Auditing of death certificates and/or medical records may help re-assess COVID-19 death attribution. For example, auditing efforts decreased the number of COVID-19 deaths in Alameda County and in Santa Clara county by about a quarter compared with those that had been originally reported^10,11^.

Here, we address these issues in a community that witnessed the first major SARS-CoV-2 outbreak in Germany, as carnival festivities around February 15, 2020 were followed by a massive outbreak. To investigate this outbreak, we previously reported a seroepidemiological observational study that found a modest IFR^12^. However, questions emerged whether early during the pandemic COVID-19-related deaths may have been missed, mislabeled or underestimated. Therefore, in the currently presented work we analyzed deaths in the studied community over time and audited death certificates during the early phase of the pandemic. Specifically, we requested official death certificates of individuals who died in the community between March and October of 2020 and analyzed in detail the underlying causes for their deaths. Furthermore, we also assessed seroprevalence with repeated surveys over a year and specifically assessed seroreversion. These analyses allow obtaining better insights on IFR estimates and on the impact of the pandemic in this intensively-studied community.

## Methods

### Death certificate assessment

Between March and October 2020 104 inhabitants of the studied community (n=12,597) deceased according to information we obtained from the state office for information and technology in North Rhine-Westphalia (Germany). With permission by the government of North Rhine-Westphalia (Germany), the state attorneys provided us with the death certificates of 77 subjects. Out of the remaining 27 deaths none was attributed to COVID-19. All death certificates were assessed by three of the authors (E.R., D.L., and H.S.) and consensus was achieved by applying the following procedures: We extracted information about gender, date of birth, date of death, COD, comorbidities and any SARS-CoV-2 infection on record with the corresponding date of the first positive test. We classified causes of deaths in the following categories: respiratory failure; sepsis; cardiovascular disease (CVD); liver failure; kidney failure; cancer; other as well as comorbidities in the categories: respiratory; sepsis; CVD; liver malfunction; kidney malfunction; cancer; neurological; diabetes; unknown; other. We also compared the number of deaths attributed to COVID-19 in the data submitted by the authorities to the national death counts versus the number of deaths attributed to COVID-19 after our audit. In attributing deaths to COVID-19 in our audit of death certificates, we followed the guidance of the WHO guidelines. Accordingly, a COVID-19-related death is defined as resulting from a clinically compatible illness in a probable or confirmed COVID-19 case, unless there is a clear alternative COD that cannot be related to COVID disease (e.g. trauma) and a death due to COVID-19 may not be attributed to another disease (e.g. cancer). In addition, there should be no period of complete recovery from COVID-19 between illness and death^13^. We could not obtain medical records for the deceased for further in-depth audit of these records.

We also retrieved information on the number of individuals in the community who live in institutionalized settings (nursing homes). It was not possible to retrieve separate counts for deaths occurring among nursing home residents either for all deaths or for COVID-19 deaths specifically.

### Seroprevalence survey in repeated visits

919 study participants, which we already enrolled in our previous study in April 2020^12^ were contacted in October 2020 (Visit 1), January 2021 (Visit 2) and April 2021 (Visit 3) by letter and were invited to an study acquisition center. After having provided written and informed consent, study participants completed a questionnaire querying demographics, symptoms, underlying diseases and medication, as well as their SARS-CoV-2 vaccination status. For children under 18 years, written and informed consent was provided by the persons with care and custody of the children following aged-adapted participant information. The 411 people (n=297 of them from this specific community) who had participated in the main carnival session “Kappensitzung”^14^ were enrolled in a separate study. Furthermore, study participants were asked to provide blood specimens and pharyngeal swabs. Over the course of all three follow-up visits, in total 587 participants from the originally study participated and provided blood specimens as well as pharyngeal swabs. Total participants for each visit are: Visit (1) 587; Visit (2) 488 and Visit (3) 406. During all three visits participants provided blood specimens and pharyngeal swabs on site in the study acquisition center. Afterwards, samples were transported to the Institute of Virology of the UKB and the cold chain remained uninterrupted during transport. Blood was centrifuged and EDTA-plasma was stored until analysis (−80 °C). Swab samples were homogenized by short vortexing and viral RNA was extracted via the chemagic Viral 300 assay (according to manufacturer’s instructions) on the Perkin Elmer chemagic™ Prime™ instrument platform. The presence of two viral target genes (E and RdRP) was assessed in each sample by real time RT-PCR (SuperScript™III One-Step RT-PCR System with Platinum™ TaqDNA Polymerase, Thermo Fisher). The following primers were used, for E gene: E_Sarbeco_F1 and R, and probe E_Sarbeco_P1, for RdRP gene: RdRP_SARSr_F, and R, and probe RdRP_SARSr-P2. In addition, an internal control for RNA extraction, reverse transcription, and amplification was applied to each sample (innuDETECT Internal Control RNA Assay, Analytik Jena #845-ID-0007100). If amplification occurred in both virus-specific reactions samples were considered positive. Plasma was used to determine Anti-SARS-CoV-2 IgG (against S1 domain) using enzyme-linked immunosorbent assays (ELISA) on the EUROIMMUN Analyzer I platform. According to the manufacturer’s instructions a result was considered positive when a ratio (extinction of sample/extinction of calibrator) of 0.8 or higher was reached. In addition, the Roche Elecsys® N ELISA was used to determine antibody response against nucleocapsid (N) of SARS-CoV-2. The result is given in COI (cutoff index), which is positive between 1.0 and approximately 250. Further we estimate for each visit after the original one how many participants had undergone seroreversion, defined as ratio of IgG below 0.8. Finally, for each visit we report on the number and proportion of participants who had tested positive by RT-PCR and we compare this against the proportion of people in the overall community who had tested positive by RT-PCR and were thus considered to be documented cases.

### Statistical analysis

Analysis of Death certificate data: Parametric normality assumptions were not applied to avoid distributional misspecifications in moderate to small samples where a central limit theorem may not be applicable. Fisher’s exact two-sided test was used to test for independence between two factor variables with two levels (2×2 designs). Pearson’s Chi-squared test was used to test for independence between factor variables and Pearson’s’ Chi-squared goodness-of-fit test was used to test hypotheses about categorical distributions. For both chi-square tests, the *p*-values were simulated using Monte-Carlo simulations with 50,000 replications to consistently approximate the small sample distributions under the null-hypotheses^15^. The one-sided two-sample Kolmogorov-Smirnov test was applied to test for a location shift in the number of days from a positive COVID-19 test and death between individuals with or without COVID-19 underlying cause of death (COD). The direction of this one-sided test was determined by the ex-ante clinical experience that SARS-CoV-2-associated death occur relatively early after infection; this experience is confirmed by the data showing an empirical first-order stochastic dominance for the group of individuals who had no COVID-19 underlying COD. A logistic regression model is used to analyze the association between the probability of a SARS-CoV-2-associated death and the amount of days from a positive COVID-19 test until death. Missing and unknown values were not imputed since these concerned only very few data points: there were two missing values in the amount of days from positive COVID-19 test until death, and there was one missing value in the age variable. Binomial confidence intervals are computed using the Clopper-Pearson method.

Analysis of seroprevalence data and IFRs: For sample size calculations, see the previous study^12^. Infection rates obtained from IgG measurements were corrected for misclassification bias using the matrix method^16^, based on sensitivity and specificity values reported by the manufacturer ELISA (Euroimmun, Lübeck, Germany; validation data sheet - version April 7th, 2020). Confidence intervals for the infection rates were computed using a bootstrap procedure based on 10,000 bootstrap samples with clustering on household-level by resampling individuals in household clusters with replacement^17^. As the bootstrapped data do not show severe abnormalities and the sample sizes are large, we estimate confidence intervals by the 2.5th and 97.5th percentiles of the bootstrap distribution. The generated bootstrap distribution was symmetrical and close to Gaussian (as indicated by normal quantile-quantile plots, with mean 0.153 and standard deviation 0.016). To analyze seroreversion, defined as IgG values below the ratio of 0.8, we analyzed participants who were IgG(+) at baseline and participated at least in one of the three follow-up visits. Wilcoxon sign-rank tests are used to statistically evaluate the change of IgG ratio with time. Here, the resulting p-values were adjusted for multiple testing following the Bonferroni-Holm procedure. Confidence Intervals (CIs) for the IFR were computed by dividing the number of deaths by the CI limits of the estimated number of infected. We estimated IFR using different acquisition periods (7, 20, 35, and 60 days after the initial seroprevalence survey^12^) as this has been a debated issue in previous meta-analyses of IFR^18,19^. The numbers of SARS-CoV-2-associated deaths in the studied community for given lengths of acquisition periods can be considered fixed as the data acquisition corresponded to a complete survey of all recorded SARS-CoV-2-associated deaths in the community during the study period. In addition, we generated also separate IFR estimates for the ages groups 0-54, 55-74 and > 75 years. To additionally account for possible uncertainty in the number of SARS-CoV-2-associated deaths we use a Bayesian credibility interval for the IFR which, however, can only be interpreted for populations for which the full survey in the community means a representative sample. This CI was computed as the empirical 2.5% and 97.5% quantiles of 100,000 samples drawn from a a-posteriori beta distribution with parameters α = [SARS-CoV2-associated deaths] + 1 and β = [estimated number of infected] - 7 + 1. This a-posteriori distribution results from a binomial likelihood model for the number of SARS-CoV-2-associated deaths with parameters trials = [estimated number of infected] and successes = [SARS-CoV2-associated deaths], and an uninformative uniform prior distribution for the IFR. To account for uncertainty in the number of infected, we use Monte-Carlo integration by sampling the estimated number of infected, for each of the 100,000 samples, form a Gaussian distribution with mean 0.153 and standard deviation 0.016 multiplied by 12,597 (i.e. the number in inhabitants in the community, January 1, 2020).

All confidence intervals were computed using a 95% coverage probability. The statistical analysis was carried out using version 4.1.1 of the R Language for Statistical Computing (R Core Team 2021: R: A Language and Environment for Statistical Computing, R Foundation for Statistical Computing, Vienna, Austria).

### Statement of Ethics

The Ministry of Health in North Rhine-Westphalia (Germany) approved the request for access to the death certificates and the district attorney of every city where the patients died provided the certificate. The original study was approved by the Ethics Committee of the Medical Faculty of the University of Bonn (approval number 085/20) and has been registered at the German Clinical Trials Register (https://www.drks.de, identification number DRKS00021306).

## Results

### Death certificate audit for March-October 2020

In total we analyzed the death certificates of 77 individuals who died between March 7^th^ and October 9^th^ 2020 in the studied community. The median age was 82 years [range 28 – 98] and 55.8% were male. Among the 77 deaths, overall 18 individuals had been tested SARS-CoV-2 positive, of whom 61.1% were male. Among the remaining 59 individuals we screened for potential signs of COVID-19 infection but no case of acute respiratory disease (ARDS) or similar cause of deaths that may be related to COVID-19 was found, thus we excluded the possibility of an undiagnosed SARS-CoV-2 infection. For the 18 individuals who had tested positive for SARS-CoV-2, we analyzed the date of death as well as the main COD (**Table 1**). 10 out of 18 individuals died in March and April 2020, seven individuals between May and August 2020, and one individual in October 2020. Deaths per month and age stratum from the 77 deaths we were able to examine are shown in **Supplemental Table 1** and they suggest a possible clustering of deaths in March 2020 only in the >70 years stratum. A total of 236 people (1.9% of the community population) lived in nursing homes during 2020 (80 male, 156 female).

**Table 1:** All individuals who were SARS-CoV-2 positive and died on COVID-19 underlying and non-COVID-19 underlying causes of death in the community. Note that patient 14 was fully recovered from COIVD-19.

| PatID | Days between positive PCR test and death | Cause of death |
| --- | --- | --- |
| *SARS-CoV-2 positive, COVID-19 underlying cause of death* |  |  |
| 1 | 1 | hypoxic lung failure, coronary heart disease |
| 2 | 5 | hypoxic lung failure |
| 3 | 0 | pneumonia |
| 4 | 3 | acute respiratory distress syndrome (ARDS), pneumonia |
| 5 | unknown | ARDS, septic shock, heart failure |
| 6 | 7 | pneumonia, multiple organ failure |
| 7 | 6 | multiple organ failure |
| 8 | 24 | circulatory failure, septic shock |
| 9 | 40 | ARDS, pneumonia |
| 10 | unknown | global respiratory failure, pneumonia |
| 11 | 32 | pneumonia, multiple organ failure, septic shock, |
| 12 | 62 | global respiratory failure, pneumonia |
| *SARS-CoV-2 positive, different underlying cause of death* |  |  |
| 13 | 24 | acute kidney failure, lactic acidosis (diabetes type II) |
| 14 | 103 | cardiovascular failure, respiratory failure |
| 15 | 92 | acute kidney failure (exsiccosis) |
| 16 | 122 | liver and kidney failure |
| 17 | 156 | oesophageal cancer |
| 18 | 187 | decompensated cardiac insufficiency |

We determined for 12 out of 18 deaths (67% (95% CI: [41%, 87%])) with cases of ARDS and respiratory failure, pneumonia, septic shock and multi organ failure as COD. The death certificates of these 12 patients stated they were admitted to the hospital with an acute SARS-CoV-2 infection, where their state of health deteriorated rapidly. Most of the cases were admitted with already advanced pneumonia, which developed into ARDS with subsequent multiple organ failure or septic shock as additional COD. Conversely, in six out of 18 deaths (33% (95% CI: [13%, 59%])) with confirmed SARS-CoV-2 infection we identified other factors to be the underlying COD. Two individuals died as a result of acute kidney failure after lactic acidosis and exsiccosis. They both had a medical history of diabetes mellitus type II, chronic obstructive pulmonary disease (COPD) and dementia, and in both cases their state of health deteriorated rapidly due to the underlying diabetes mellitus. Additionally, they had refused to eat and drink as a result of dementia, which ultimately led to acute kidney failure and exsiccosis. One individual had decompensated cardiac insufficiency with medical history of food refusal, COPD, epilepsy and Korsakov’s disease, while another had a history of advanced and metastasized esophageal carcinoma. The death certificates of these cases described a progressively deteriorating general condition of the patient, with eventual death from their underlying illnesses. Lastly, there was one case of acute liver and kidney failure as stated COD with no documented comorbidity on the death certificate and one case of cardiovascular failure as well as respiratory insufficiency with medical history of diabetes and cardiovascular disease. In the latter case it was noted, that the individual had previously recovered from SARS-CoV-2 infection. In addition, five out of the six individuals died three to six months after confirmed SARS-CoV-2 infection and more importantly the death certificates of all six cases reported recovery from COVID-19, which imply a period of complete recovery from COVID-19 between illness and death. Therefore, these cases do not fulfill the criteria of the WHO guidelines to be counted as COVID-19 deaths. Overall, only 12 out of 18 deaths with confirmed SARS-CoV-2 infection died because of COVID-19 between March and October 2020. During the same time period, the authorities had reported 15 COVID-19 deaths to the national COVID-19 death counts^20^. Therefore, apparently 3 of the deaths that were not caused by COVID-19 had been officially reported as COVID-19 deaths.

Next, we analyzed differences in age, COD and comorbidities among all deaths that had occurred in the studied community. We therefore grouped the 77 deaths in three mutually exclusive groups (SARS-CoV-2 negative; SARS-CoV-2 positive and COVID-19 underlying COD; SARS-CoV-2 positive but no COVID-19 underlying COD). There were no statistically significant differences in age distribution (median 84.5 [28-98], 80 [56-86], 75 [56-82], respectively, p=0.1597) (**Figure 1**) between these groups. However, while individuals with confirmed SARS-CoV-2 infection and no SARS-CoV-2-associated death had no common COD, individuals with confirmed SARS-CoV-2 infection were more likely to have died from respiratory failure compared to other COD (sepsis and CVD) (75% versus 25%, p=0.01). Moreover, only 8% of the individuals without COVID-19 underlying COD had no comorbidity listed on the death certificate, whereas this was the case for 33% of individuals with COVID-19 underlying COD (p = 0.029).

**Figure 1:**
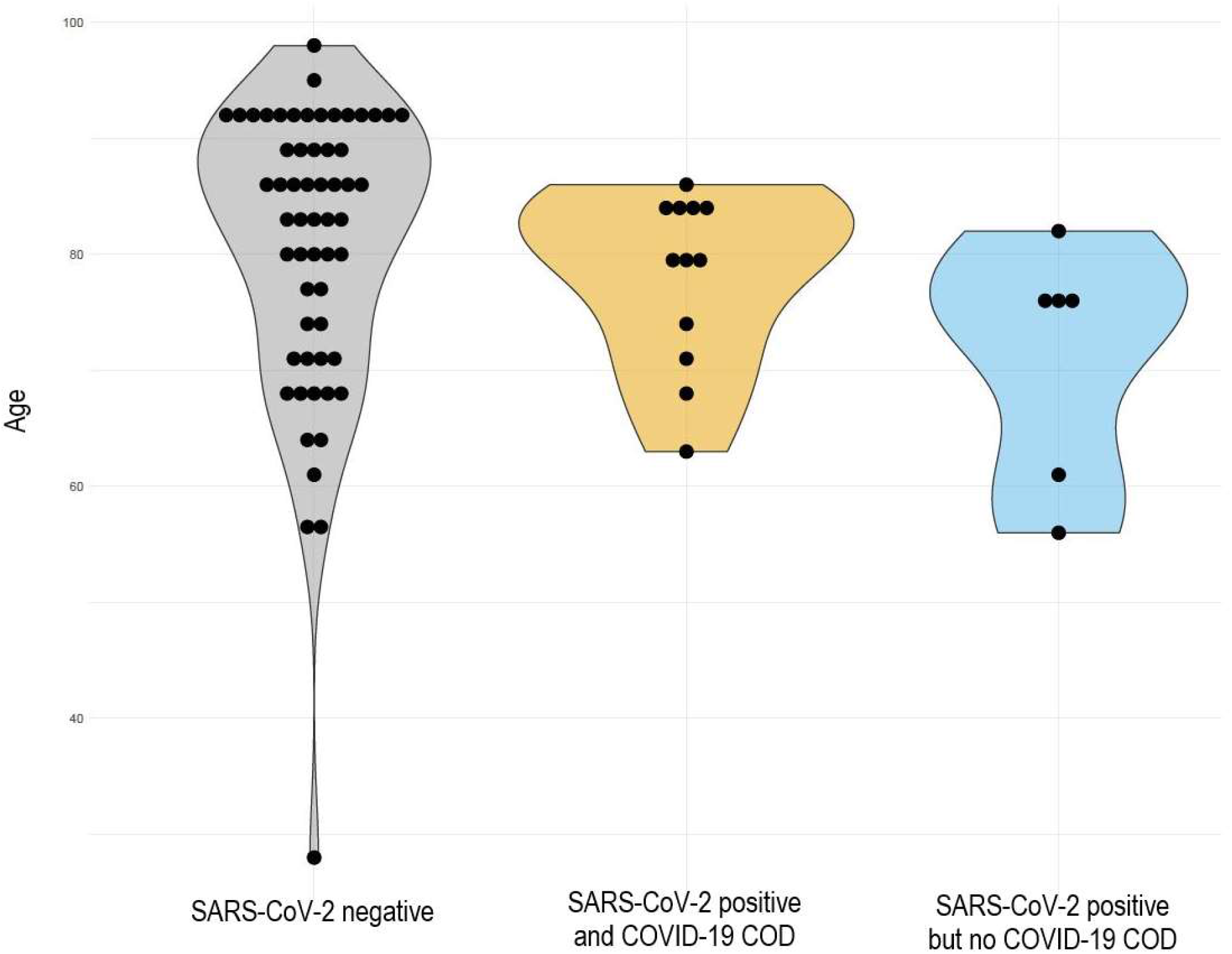
Age differences. All 77 deaths were differentiated in three groups: SARS-CoV-2 negative, SARS-CoV-2 positive and COVID-19 underlying COD as well as SARS-CoV-2 positive but no COVID-19 underlying COD. There were no significant differences in the age distribution among COVID-19 infected or non-infected individuals.

In individuals with SARS-CoV-2-associated deaths a median of 18 days [0 – 62] passed between positive PCR-test and death, a shorter time span compared to individuals with SARS-CoV-2 infection but not COVID-19 underlying COD (median 144 days [24 – 187]) (**Figure 2**). The number of days from positive COVID-19 test to death were higher (p=0.005) for individuals who had no SARS-CoV-2-associated death. In logistic regression, the probability to die of COVID-19 decreases (p=0.04) for larger numbers of survived days after a positive COVID-19 test. Overall, the likelihood to die of COVID-19 decreased markedly 100 days or more after confirmed SARS-CoV-2 infection (**Supplemental Figure 1**).

**Figure 2:**
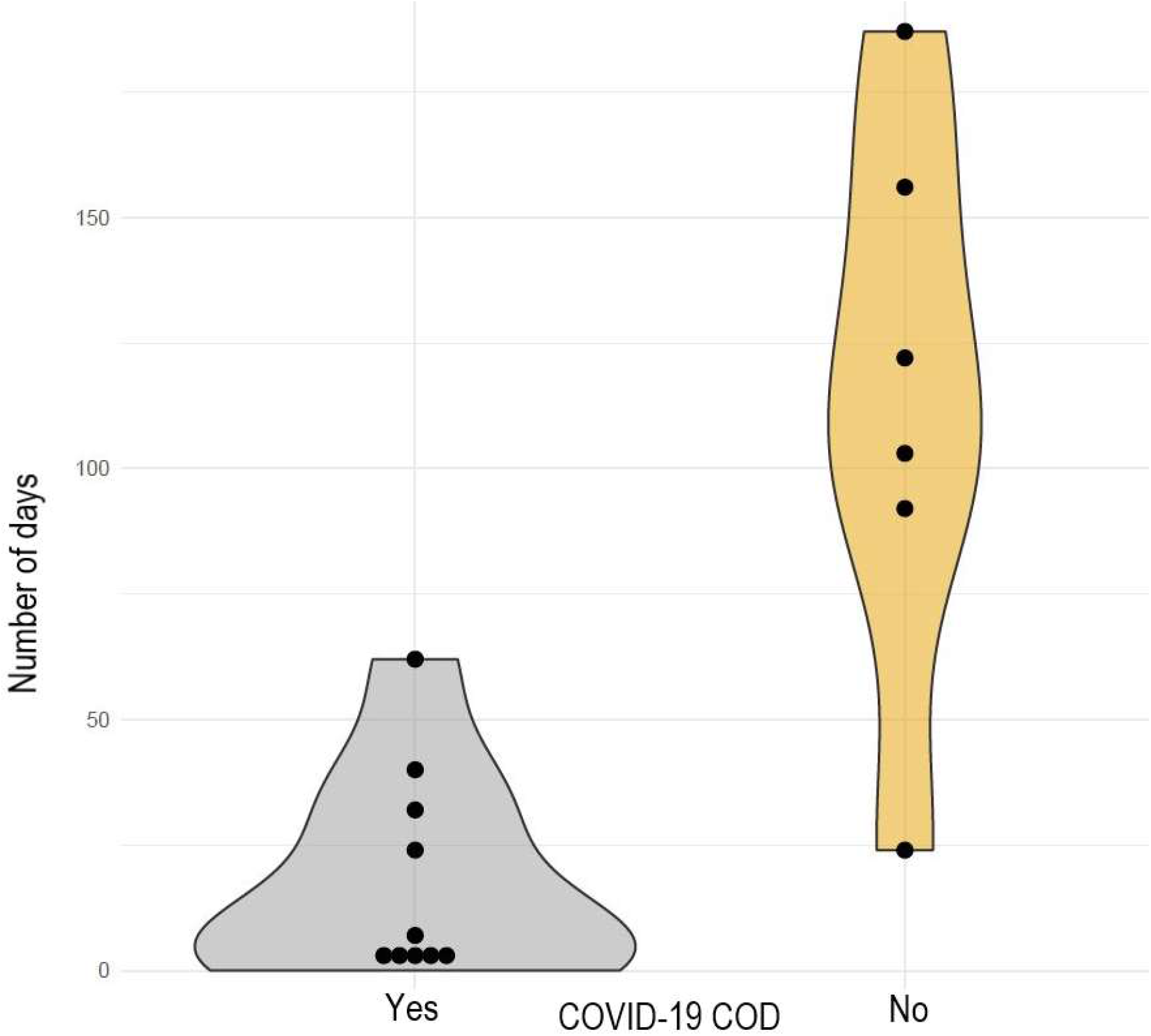
Number of days between confirmed SARS-CoV-2 PCR test and death of the individual. Stratified by COVID19 underlying cause of death. Individuals with SARS-CoV-2-associated deaths had with a median of 18 days [range 0 – 62] between positive PCR-test and death, a shorter time span compared to individuals with SARS-CoV-2 infection but not COVID-19 underlying COD with a median of 144 days [range 24 – 187].

### Infection fatality rate

Figure 3. shows the estimated IFR with different acquisition periods ranging from 7 days (7 deaths), 20 days (8 deaths), 35 days (9 deaths), and 60 days (12 deaths). Extending the observation period further did not lead to a change in the IFR depending on the acquisition period. The IFR estimates ranged accordingly from 0.36% to 0.62%, the 95% confidence intervals ranged from 0.28% to 0.84% and this confidence range would become 0.18% to 1.15% when introducing also uncertainty in the number of deaths. **Table 2** shows the IFR estimates for age strata 0-50, 55-74, and >75 years. As no deaths occurred in the age stratum 0-50, the IFR estimates are all 0.0% For the 55-74 years the IFR estimates range from 0.18 to 0.74 and the 95% confidence intervals extend from 0.13 to 1.19 For people >75 years old, the IFR ranges from 3.21 to 4.28 and the 95% confidence intervals extend from 1.87 to 9.21.

**Table 2.** Estimates of infection fatality rate and 95% confidence intervals in age strata with different acquisition windows.

|  | 0 – 54 years | 55 - 74 years | >75 years |
| --- | --- | --- | --- |
| **Acquisition period** |  |  |  |
| 7 days acquisition period | 0 [0.0] | 0.18 [0.13, 0.30] | 3.21 [1.87, 6.91] |
| 20 days acquisition period | 0 [0.0] | 0.18 [0.13, 0.30] | 3.75 [2.18, 8.06] |
| 35 days acquisition period | 0 [0.0] | 0.37 [0.25, 0.60] | 3.75 [2.18, 8.06] |
| 60+ days acquisition period | 0 [0.0] | 0.74 [0.50, 1.19] | 4.28 [2.50, 9.21] |

**Figure 3:**
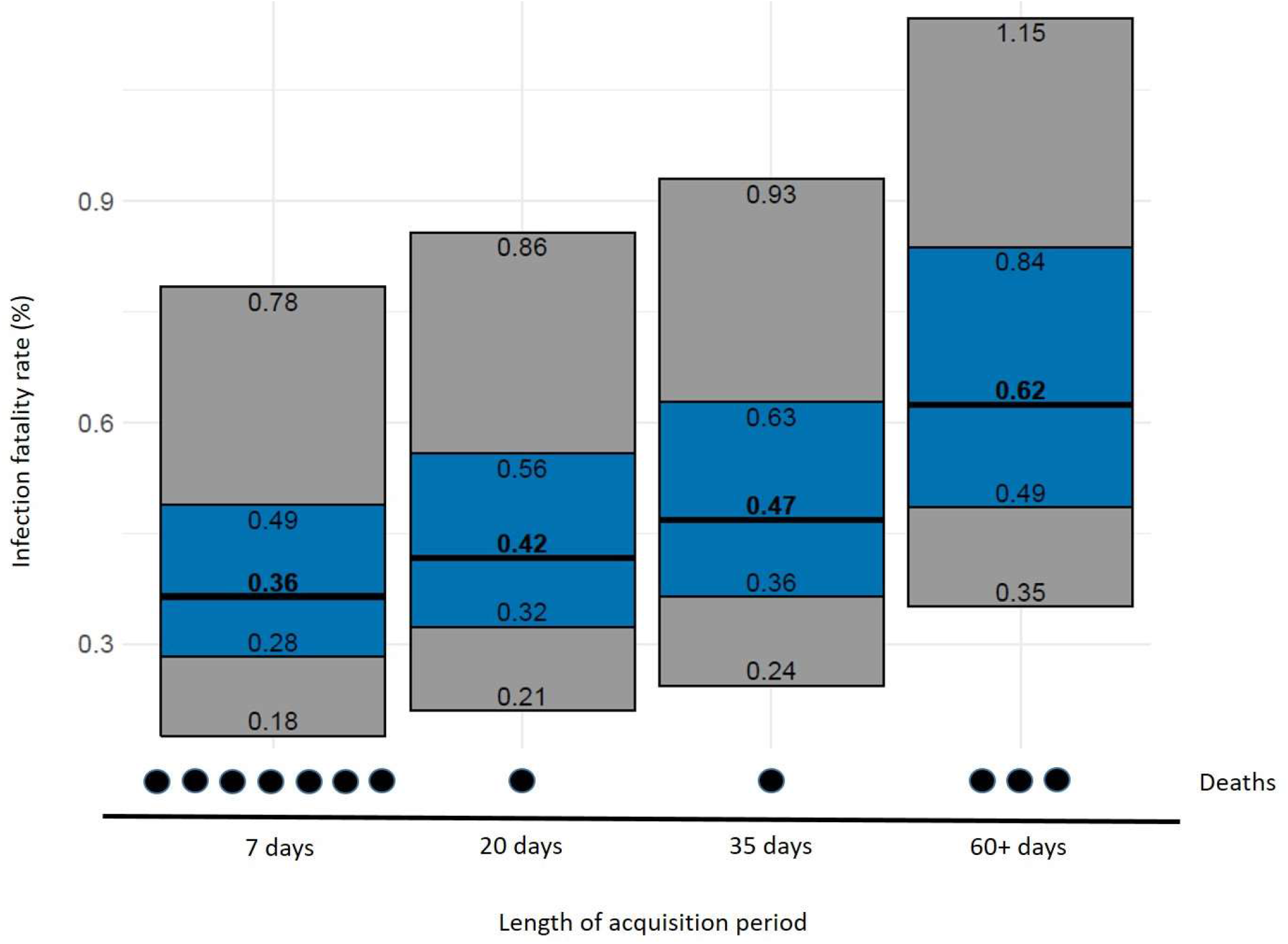
Newly calculated infection fatality rate (IFR) with 95% confidence intervals (blue) and 95% credibility intervals (gray) additionally accounting for uncertainty in the number of deaths.

### Repeated seroprevalence surveys

While 18.1% (95% CI: [13.7%, 23.0%]) of all study participants were found to be IgG(+) 6 months after the original study was conducted (baseline), this number increased 9 and 12 months later (21.0% [16.2%, 26.1%]; 35.9% [30.0, 42.0] respectively). Values were corrected for sensitivity and specificity of IgG (sensitivity 90.9%; specificity 99.1%). Next, we wanted to understand how many participants had undergone seroreversion, defined as IgG values below the ratio of 0.8. Therefore, we analyzed participants who were IgG(+) at baseline and participated at least in one of the three follow-up visits. We could see differences in the IgG levels of the participants already 6 months after the initial visit. IgG was significantly decreased over time and showed reduced antibody titers at 6 and 9 months (p<0.0001) (**Figure 4**). In total 27 (30%) out of 90 participants (95% CI: [20.5%; 39.5%]) who were IgG(+) at time point 0 seroreversed and showed no detectable IgG levels after 12 months. Finally, we analyzed the pharyngeal swabs via RT-PCR to identify active SARS-CoV-2 infection. Overall, only 2 participants were tested positive in our cohort over the 12 months of follow-up. One participant at 6 months, who was IgG(+) at baseline but had no detectable IgG antibodies 6 months later. The second participant was RT-PCR(+) at 9 months and was IgG negative at 0 and 6 months. More importantly, the proportion with positive PCR in our seroprevalence-tested cohort was far smaller (2 out of 587 tested participants, i.e. 0.3%) than the respective proportion in the overall population of the community (n=374 PCR confirmed cases, i.e. 3.0%) in the same period of time, between October 2020 and April 2021.

**Figure 4:**
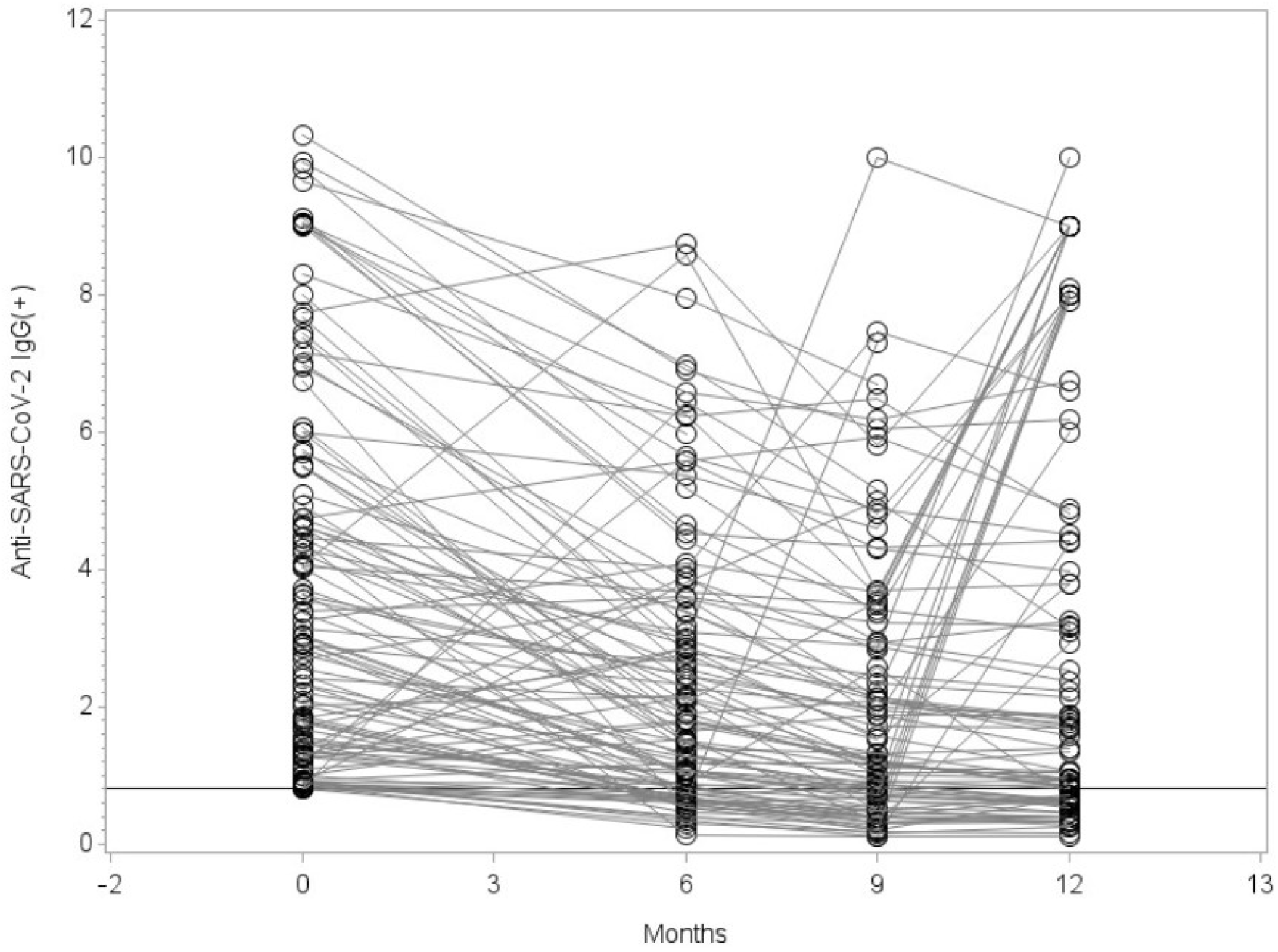
IgG levels over time of participants who were IgG(+) at baseline and participated at least in one of the three follow-up visits were analyzed. 27 participants who developed IgG values below the ratio of 0.8 (Limit of Quantification) were considered as seroreversed. Statistical significance was assessed using Wilcoxon sign-rank test and the resulting six p-values were adjusted for multiple testing following the Bonferroni-Holm procedure. Visit 1 vs. 0: p<0.0001; Visit 2 vs. 0: p<0.0001; Visit 3 vs. 0: p=0.0016; Visit 1 vs. 2: p<0.0001; Visit 1 vs. 3: p=0.0053; Visit 2 vs. 3: p<0.0001. All comparisons show a decreasing trend with time. It is important to note, that although the graph only shows those with IgG ratio ≥ 0.8 at visit 0, all subjects who contribute data for visit 0 and at least one of the following visits are included in the test (without restriction at visit 0).

## Discussion

In this study, we audited and analyzed the deaths occurring in a German community between March and October of 2020 at the beginning of the SARS-CoV-2 pandemic. In total 18 dying individuals were reported positive for SARS-CoV-2, but COVID-19 was the underlying COD in only 12 out of the 18 deaths with confirmed SARS-CoV-2 infection. Therefore, one third of individuals with confirmed SARS-CoV-2 infection died with non-COVID-19 related COD. The duration between confirmed SARS-CoV-2 infection and death was significantly higher in individuals without COVID-19 related COD compared to those with COVID-19 related COD. Moreover, the number of deaths reported to national authorities as COVID-19 deaths was 25% higher than the truly “causal” COVID-19 deaths. This over-estimation of COVID-19 deaths is similar to the rates seen in audits in US counties^10,11^. Over-estimation of COVID-19 deaths may apply also to other settings, especially in high-income countries where intensive testing is performed^9^.

We also observed that more SARS-CoV-2 positive individuals with non-COVID-19 underlying COD had listed comorbidities on the death certificates compared to individuals with COVID-19 underlying COD. This finding needs to be interpreted with caution given the small numbers and the possibility that comorbidities are often not fully reported in death certificates. Unfortunately, we could not have access to the full medical records. Thus we cannot exclude the possibility that some of the certificates where the COD was COVID-19 simply failed to report also existing co-morbidities.

The published WHO guidelines define a COVID-19-associated death as a death resulting from a clinically compatible illness, in a probable or confirmed COVID-19 case, unless there is a clear alternative cause of death^13^. Our analysis of death certificates underlines the importance to more accurately confirm COD of individuals who have died with a confirmed SARS-CoV-2 infection, especially when the elapsed time between first SARS-CoV-2 positive test and death is long. We could show that 33% with confirmed SARS-CoV-2 infection had other factors to be the underlying COD, and taking into account the CI of (95% CI: [13%, 59%]) this is coherent with the recently published cause-of-death statistics of 2020 for Germany from the Federal Statistical Office^18^. Recent publications have highlighted difficulties in counting deaths with COVID-19 underlying COD and the need of a continuous surveillance of mortality records^19,21^. Reported studies of COVID-19-associated deaths in Germany and the UK determined septic shock, multi organ failure and respiratory failure as the most common immediate COVID-19 underlying COD, often due to suppurative pulmonary infection and diffuse alveolar damage^22-24^. Our findings for our studied community are consistent with this, as 75% of the COVID-19-associated deaths died from respiratory failure in combination with septic shock and multi organ failure. Furthermore, it was described that smoking or some comorbidities like cancer and chronic liver disease had stronger associations with non-COVID than COVID-19 deaths in association with age^22^. Although we do not see significant associations with age or specific comorbidities (probably due to the low number of cases), we determined kidney failure, lactic acidosis or cancer as the main causes in non-COVID-19 associated deaths. These findings support the importance of looking more carefully into the deaths of individuals with a confirmed SARS-CoV-2 infection and taking into account the individuals’ medical history along with their most recent medical data and symptoms.

Recent publications on COVID-19 disease progression analyzed the length of stay in the hospital and survival time^25-27^. It was reported that the median length between symptom onset and hospitalization ranged between 3 and 10.4 days, depending on the age of the patient^26^. However, length of stay in hospital for patients who died eventually were an additional six to seven days as reported in a Belgian study^26^, and for China^27^ the estimated mean time from symptom onset to death was 18.8 days. Although a positive PCR test for SARS-CoV-2 does not necessarily coincide with the onset of symptoms, the results are consistent with our findings. Individuals in our studied community with SARS-CoV-2-associated deaths had a median of 18 days between positive PCR-test and death, whereas individuals with no COVID 19-underlying COD had a longer time span with a median of 144 days. The likelihood to die of COVID-19 decreased sharply with longer follow-up markedly after confirmed SARS-CoV-2 infection.

Evaluation of the deaths per month and age stratum showed a peak of deaths in March 2020 and it was concentrated in people over 70, consistent with the age of the early COVID-19 fatalities. We could not retrieve information on how many of these individuals might have been nursing home residents. However, this specific community has a high proportion of its population residing in nursing homes (about 2 times larger than the average for Germany [731,000 people, 0.9% of the German population])^28^. It is possible that many of the COVID-19 fatalities in the community might have been in people with limited life expectancy, regardless of whether they were institutionalized or not. Obviously, one has to be cautious with these inferences, especially given the relatively small number of deaths. Of note, eventually the overall number of deaths in the community for calendar year 2020 (n=157) was similar to calendar years 2018 (n=156) and 2015 (n=158) but higher than other previous recent years. However, it is very likely that this was influenced by the hard lockdown in the studied community in March and April 2020.

Furthermore, repeated seroprevalence surveys showed that seroprevalence changed only modestly in the fall of 2020 and beyond as compared with our previously published results on seroprevalence in April 2020^12^. However, we documented substantial rates of seroreversion, which is in agreement with some other studies^29-36^. Some studies have found even higher rates of seroreversion, e.g. one investigation found a median time to seroreversion for IgG being only 55 days^30^, but this may include false-positives at initial screening. Nevertheless, it is very likely that seroreversion causes the number of infected individuals to be substantially under-estimated and thus the IFR to be over-estimated^29,30^. This may apply even to our early April 2020 survey that happened within two months of the superspreader event.

Estimates of number of people infected in the population may also be affected by the representativeness of the surveyed sample. In our original seroprevalence survey in April 2020^12^, we had noticed that the tested sample had a lower proportion of documented RT-PCR infections than the overall community population (2.39% versus 3.08%). Correction for this factor would decrease IFR estimates by 29% (e.g. from 0.36-0.62% to 0.28-0.48%). The sampling deviation was seen also, even more prominently in the follow-up surveys, where only 2 participants tested positive by RT-PCR (10-fold less than in the general population). It may reflect the fact that participants (especially those who return also for follow-up visits) may be more health conscious; or alternatively, less likely to perform RT-PCR testing since they are tested for antibodies. We also observed that our original seroprevalence study sample was under-representative of individuals infected during the main carnival event, the Kappensitzung, where participants had very high infection rates^14^, three times higher than the general population of the community by April 2020. Due to the small number of cases, we were not able to exclude that this may have been entirely due to chance but the pattern is consistent with the possibility that seroprevalence is under-estimated (and, correspondingly, IFR over-estimated). These observations highlight the difficulty of using seroprevalence samples and repeated surveys to assess the number of infected individuals in a population.

Our observations show that the calculation of IFRs needs to be done very carefully and that their interpretations need to take into account all possible influencing factors. In our study, over-estimation of COVID-19 deaths and under-estimation of seroprevalence may have inflated the IFR estimate. Additional variability in the estimates may be introduced by the time window used for capturing deaths, its relationship to the time window of the seroprevalence, and the assumptions made about the delay in developing antibodies and in dying after infection. In the case of this specific community, these uncertainties would probably still be captured by the original confidence interval that we reported for IFR in our original publication and that allowed for uncertainty in the number of deaths (95% CI, 0.17% to 0.77%). Moreover, the exact case mix of infected individuals can have a major impact on the IFR, given the extremely steep age gradient that has been documented before and which we also saw prominently in the community data^5,37^. It should be noted that due to the study-design, the results of this study can only be representative for this specific community. Overall, in seroprevalence studies where only a tiny portion of the population is selected for serological testing (e.g. typically 0.01-1% in nation-wide surveys^38^) one should remain cautious, as errors in counting deaths, seroprevalence, and other sources of uncertainty may have an larger impact.

In conclusion, our in-depth assessment of the fatality impact of COVID-19 in the community shows a relatively low fatality rate in this community with deaths concentrated entirely in the elderly, but seroprevalence estimates that go into IFR calculations need careful considerations of their time window, potential seroreversion, and representativeness. Importantly, COVID-19 deaths may have been overcounted and the relative contribution of over- and under-counting of COVID-19 deaths needs careful auditing across multiple other locations.

## Data Availability

All data produced in the present study are available upon reasonable request to the authors.

## Acknowledgments

We would like to thank all the subjects who participated in our repeated seroprevalence surveys. Furthermore, we would like to thank the following people who helped with the study: Maximilian Baum, Celina Beta Schlüter, Melanie Geiger, Annika Breuer, Julia König, Karola Mai, Antonia Büning, Paulina Tarnow, Annina Hahn, Désirée Deloud, Anna-Lena Suchan, Sven Kohrn, Monika Eschbach-Bludau, Tobial Höller as well as the local government and physicians for their support to conduct the study.

## Author contribution

E.R., D.L., J.P.A.I., and H.S. contributed to the conception, design, and interpretation of the work. E.R., D.L., B.S., N.L., T.H., C.F., J.P.A.I. and H.S. contributed to the data acquisition and analysis of the data. K.H.J. revised the work critically for important intellectual content. E.R., D.L., J.P.A.I., and H.S. wrote the manuscript.

