## Supplemental Table and Figure for "Analysis of Fatality Impact and Seroprevalence Surveys in a Community Sustaining a SARS-CoV-2 Superspreading Event"

### Supplemental Materials

**Supplemental Table 1:** Deaths per month and age stratum from the 77 deaths we were able to examine.

| Month (2020) | Available death certificates | Age ≤ 70 | Age >70 |
|---|---|---|---|
| March | 20 | 3 | 17 |
| April | 12 | 2 | 10 |
| May | 10 | 3 | 7 |
| June | 9 | 0 | 9 |
| July | 9 | 4 | 5 |
| August | 12 | 2 | 10 |
| September | 2 | 2 | 0 |
| October | 3 | 1 | 2 |

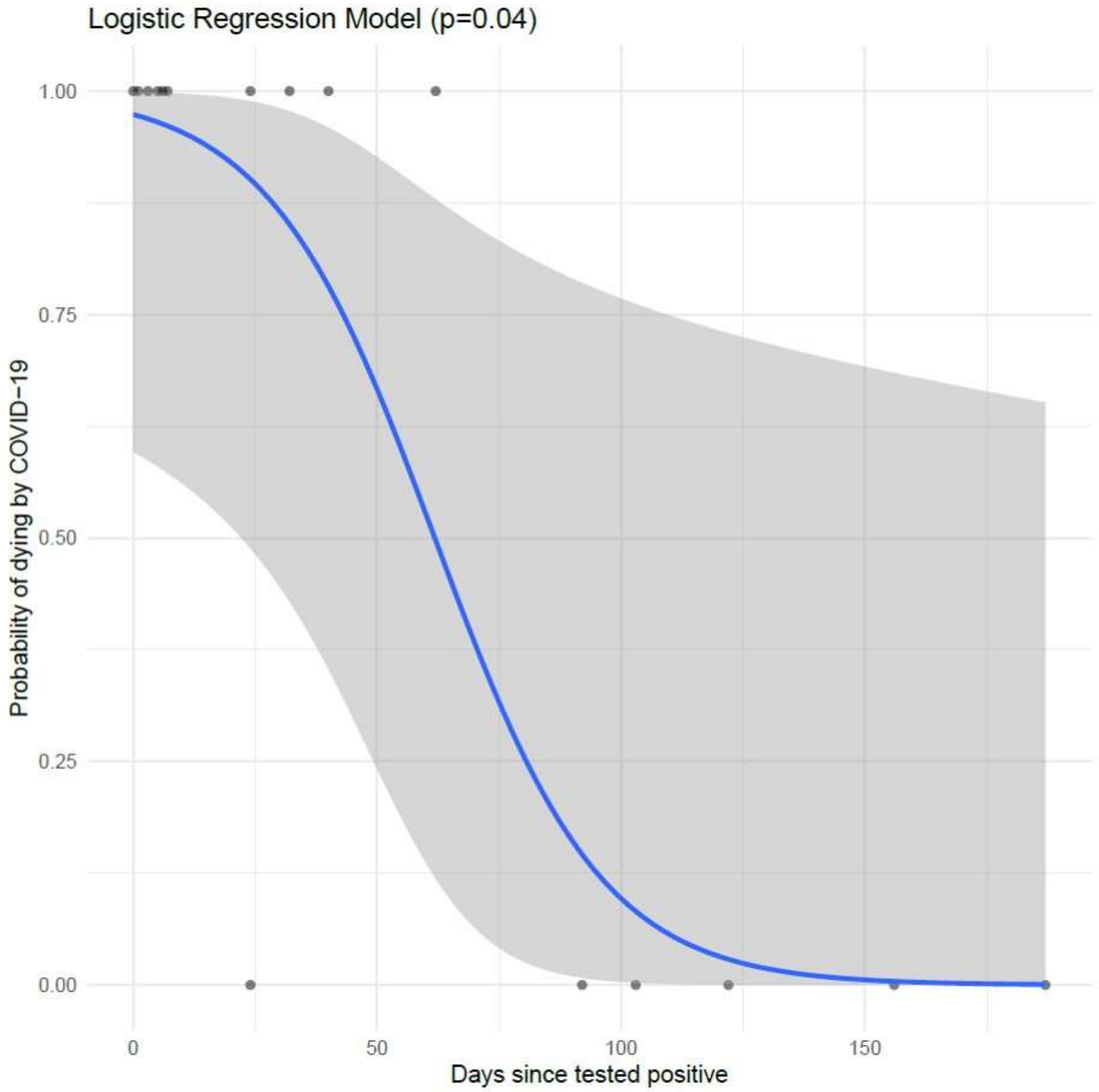


**Supplemental Figure 1:** The likelihood to die at COVID-19 underlying causes of death after positive PCR-test for severely ill individuals in the specific community we examined. A logistic progression model demonstrating that the likelihood to die of COVID-19 is significantly decreased (p=0.04), if death occurred 100 days or later after confirmation of SARS-CoV-2 infection.
